# Stress-Related Methylation Risk Scores Predict Coronary Heart Disease

**DOI:** 10.64898/2026.07.31.26359423

**Authors:** Sofia Benavides, Hazel Milla, Helena Palma-Gudiel, David Checknita, Bjoernar Tuftin, Kai Xia, Charles Kooperberg, Alexander P. Reiner, JoAnn E. Manson, Themistocles L. Assimes, Parveen Bhatti, Kent D. Taylor, W. Craig Johnson, Stephen S. Rich, Jerome I. Rotter, Linda C. Gallo, David R. Rubinow, Elior Rahmani, Laura M. Raffield, Eric A. Whitsel, Anthony S. Zannas

## Abstract

**Background:** Psychosocial stress is a key risk factor for coronary heart disease (CHD), particularly in postmenopausal women who face both a high stress burden and elevated cardiovascular risk. DNA methylation (DNAm) – a critical epigenetic modification bridging environment and health – remains understudied as a contributor to stress-related CHD.

**Methods:** We conducted an epigenome-wide association study (EWAS) of stress in the Women’s Health Initiative (WHI), an ancestrally diverse cohort of postmenopausal women (n=3,857). At screening visit, participants completed a questionnaire assessing stressful life events and provided whole blood for DNAm. Incident CHD was then longitudinally ascertained (follow-up mean/SD: 16.7/8.4 years), and DNAm signatures were evaluated as CHD predictors using Cox regression. Predictive models were independently validated in the Jackson Heart Study (JHS; n=3,053) and Multi-Ethnic Study of Atherosclerosis (MESA; n=870). The bulk-level DNAm associations were computationally deconvolved at the cell-type-specific level using tensor composition analysis (TCA).

**Results:** The EWAS in WHI identified 841 stress-related DNAm sites (99 hypermethylated, 742 hypomethylated with stress) after FDR correction, with 13 significant after Bonferroni correction, including sites located on immune and CHD-related genes (e.g., *TNF*, *ALDH2*). Methylation risk scores (MRSs) integrating the 841 FDR-significant sites (MRS_841_) and 13 Bonferroni-significant sites (MRS_13_) predicted incident CHD (HR=1.33-1.37; p≤0.0008) and mediated 16.5-17.7% of the association between stress and CHD. In JHS and MESA, MRS_13_ independently predicted CHD (HR=1.34; p=0.036), whereas MRS_841_ was suggestively associated with CHD (HR=1.27; p=0.087). TCA indicated that the greatest number of stress-related sites predictive of CHD was specifically in monocytes (133 total), with directions consistent with bulk-level associations (9 hypermethylated, 124 hypomethylated with stress).

**Conclusion:** Our study supports methylation risk scores as novel biomarkers of stress-related CHD and uncovers epigenetic regulation in monocytes as a potential underlying mechanism. These findings highlight biological pathways linking stress and disease and may promote personalized interventions in high-risk populations.

## Introduction

Despite advances in cardiovascular prevention and treatment, coronary heart disease (CHD) remains the leading cause of death in the United States^1–4^. A major challenge in alleviating CHD burden is its complex pathogenesis involving genome-environment interactions, the mechanistic understanding of which is limited^5–7^. Among key environmental factors, psychosocial stress has been implicated as a risk factor for CHD^8–14^. The role of stress in CHD is particularly relevant in postmenopausal women, who experience high stress burden relative to men and premenopausal women^15–17^ and elevated cardiovascular disease risk^18–20^. Indeed, prior work has associated higher stress burden with increased CHD risk in postmenopausal women^11,21^, which is consistent with similar observations in other populations^22–24^. Despite these observations, the molecular mechanisms linking stress and CHD remain poorly understood.

Epigenetic changes, chemical modifications that regulate gene expression without changing the underlying DNA sequence, are thought to mediate the association between environmental exposures such as stress and disease outcomes, including cardiovascular disease (CVD)^21–23^. Studies by our group and others have shown that differences in DNA methylation (DNAm), a well-established epigenetic modification to CpG sites (cytosine-guanine dinucleotides), are associated with psychosocial stress, immune dysregulation, and CVD. Our work further suggests that stress-driven DNAm changes occur at select genomic sites in peripheral blood cells and can promote inflammatory signaling associated with CHD^25^. However, the extent to which stress-related epigenetic signatures occurring across the genome predict CHD outcomes remains unclear. Moreover, whether epigenetic patterns in distinct immune cell types underlie the association between stress and CHD risk is unknown.

To this end, we leveraged large-scale longitudinal cohorts participating in the NHLBI Trans-Omics for Precision Medicine (TOPMed) program^26^. To identify stress-related DNAm sites, our discovery analyses focused on the Women’s Health Initiative (WHI), an ancestrally diverse cohort of postmenopausal women^27^, in which higher stress burden was previously shown to predict incident CHD^11,21^. To capture the overall impact of stress on the epigenome, we calculated methylation risk scores (MRSs) for each participant based on DNAm levels of stress-related CpG sites and assessed whether these MRSs predict and mediate CHD outcomes in the WHI. We independently evaluated the generalizability of these markers in two diverse TOPMed cohorts of pre- and postmenopausal women and men, the Jackson Heart Study (JHS) and Multi-Ethnic Study of Atherosclerosis (MESA). Because DNAm levels vary by cell type and both stress and CHD have been associated with epigenetic changes in distinct cell types^28,29^, we also implemented cell-type-specific deconvolution to identify distinct cell types underlying MRS-CHD associations.

## Methods

The code used for all analyses are available on Github (https://github.com/hazelmilla/MRS_TOPMed.git). Additional data are available from the corresponding author upon reasonable request. Details for DNAm data processing across cohorts are provided in the Supplement.

### Cohorts overview

#### Women’s Health Initiative (WHI)

The WHI, which has previously been described in detail^11,27,30^, is a multi-ethnic cohort study aimed at preventing disease and disability in postmenopausal women. It consists of 161,808 women, aged 50-79 across 40 locations. For the WHI subset included in our study, whole-blood DNAm array data (∼450 K) were available for 3,857 WHI participants from the following ancillary studies: Epigenetic Mechanisms of Particulate Matter-Mediated CVD Risk (WHI-EMPC)^11^; Broad Agency Announcement, i.e., Integrative Genomics and Risk of CHD and Related Phenotypes in the WHI (WHI-BAA23)^31^; and Ancillary Study 311 (WHI-AS311)^32^ (Table 1). CHD was defined as clinical myocardial infarction (MI), definite silent MI, or death due to definite or possible CHD. Clinical MI, CHD deaths, and definite silent MI were adjudicated through medical record review using standardized criteria^11,21,33^. Any participant included in multiple ancillary studies was represented only once in the analysis. For participants with multiple DNAm measurements, a single sample was randomly selected before analysis so that each participant contributed only one DNAm profile. Furthermore, we only included participants with DNAm collected at the initial WHI screening visit. All participants provided written consent.

**Table 1.** Cohort demographics.

| Variable | WHI | JHS | MESA |
| --- | --- | --- | --- |
| **n** | 3,857 | 3,053 | 870 |
| **SLE Burden (%)** |  |  |  |
| Lower | 1,708 (44.3) | - | - |
| Higher | 2,149 (55.7) | - | - |
| **Sex (%)** |  |  |  |
| Male | 0 (0) | 1,109 (36.3) | 414 (47.6) |
| Female | 3,857 (100) | 1,944 (63.7) | 456 (52.4) |
| **Incident CHD (%)** |  |  |  |
| No | 3,090 (80.1) | 2,797 (93.6) | 814 (93.7) |
| Yes | 767 (19.9) | 190 (6.4) | 55 (6.3) |
| **Years to Follow-Up for All Participants (mean(SD))** | 18.2 (7.8) | 12.5 (3.5) | 17.2 (2.2) |
| **Years to CHD Event for CHD Cases (mean(SD))** | 16.7 (8.4) | 6.5 (3.9) | 14.6 (2.4) |
| **Age (mean(SD))** | 63.95 (7.14) | 54.5 (12.8) | 60.25 (9.8) |
| **Marital Status (%)** |  |  |  |
| Single | 1,651 (42.8) | 1,349 (44.2) | 311 (36.0) |
| Marriage-like Status | 2,187 (56.7) | 1,698 (55.6) | 553 (64.0) |
| Data Missing | 19 (0.5) | 6 (0.2) | 6 (0.7) |
| **BMI (mean(SD))** | 29.31 (6.14) | 31.9 (7.3) | 28.8 (5.2) |
| **Smoking (%)** |  |  |  |
| No | 1,953 (50.6) | 2,641 (86.5) | 748 (85.9) |
| Yes | 1,869 (48.5) | 391 (12.8) | 122 (14.0) |
| Data Missing | 35 (0.9) | 21 (0.7) | 0 (0) |
| **Alcohol (%)** |  |  |  |
| No | 959 (24.9) | 1,600 (52.4) | 202 (27.9) |
| Yes | 2,315 (60) | 1,437 (47.1) | 523 (72.1) |
| Data Missing | 583 (15.1) | 16 (0.5) | 145 (16.7) |
| **Ethnicity (%)** |  |  |  |
| Not Hispanic/Latino | 3,462 (89.8) | 3,053 (100) | - |
| Hispanic/Latino | 389 (10.1) | 0 (0) | - |
| Unknown | 6 (0.2) | 0 (0) | - |
| **Race (%)** |  |  |  |
| Chinese | - | - | 65 (7.5) |
| Asian | 76 (2.0) | 0 (0) | - |
| Black | 1,009 (26.2) | 3,053 (100) | 161 (18.5) |
| White | 2,772 (71.9) | 0 (0) | 384 (44.1) |
| Hispanic | - | - | 260 (29.9) |
| **Education (%)** |  |  |  |
| Less than high school completed | 336 (8.7) | 471 (15.5) | 142 (16.4) |
| High school diploma or equivalent | 726 (19.7) | 622 (20.4) | 161 (18.6) |
| More than high school completed | - | 1,950 (64.1) | - |
| Some College | 1,503 (39) | - | 150 (17.3) |
| College or more completed | 1,264 (32.8) | - | 415 (47.8) |
| Data Missing | 28 (0.7) | 10 (0.3) | 2 (0.2) |
| **Income (%)** |  |  |  |
| Poor (<\$25,000) | - | 354 (13.6) | - |
| < \$35,000 | 1,809 (46.9) | - | 417 (54.1) |
| Lower-middle (\$25,000–\$49,999) | - | 626 (24.1) | - |
| \$35,000 - \$49,999 | 761 (19.7) | - | 99 (12.8) |
| Upper-middle (\$50,000–\$74,999) | - | 811 (31.2) | - |
| \$50,000 - \$74,999 | 600 (15.6) | - | 158 (20.5) |
| Affluent (≥\$75,000) | - | 810 (31.1) | - |
| > \$75,000 | 456 (11.8) | - | 97 (12.6) |
| Data Missing | 231 (6.0) | 452 (14.8) | 12 (1.4) |

Stress burden was assessed using a stressful life event (SLE) questionnaire developed in the Alameda County Epidemiological Study^34^ and modified in the Beta-Blocker Heart Attack Trial for older women^13^. This questionnaire was administered during the baseline visit and prompted participants to indicate if the following events occurred within the last 12 months: (1) death of spouse, (2) death of close friend, (3) major financial problems, (4) a divorce or break up, (5) close friend divorced, (6) major conflict with children or grandchildren, (7) involved in a major accident, (8) lost job, (9) physically abused, (10) verbally abused, or (11) pet died. Participants then indicated the degree of upset for events that occurred on a scale of 1 (not very upsetting) to 3 (very upsetting). The scale ranged from 1 to 33, with a higher score corresponding to a higher SLE burden. Due to its skewed distribution, the SLE sum score was dichotomized using an approximate median split into lower (0–2) and higher (3–33) stress burden groups, in line with prior literature employing this measure^11,21,33^.

#### Jackson Heart Study (JHS)

The JHS study design and methods have been previously described in detail^35–37^. JHS is a longitudinal, community-based observational study of genetic and environmental risk factors contributing to CVD in a population of African American men and women. Consisting of 5,306 African American residents in the Jackson, Mississippi metropolitan area, the JHS includes extensive medical, family, lifestyle, and psychosocial histories alongside physical and biochemical measurements and diagnostic procedures at baseline (2000 – 2004) and at two follow-up examinations (2005 – 2008 and 2009 – 2013). Adjudicated incident CHD was defined as definite/probable MI, fatal CHD, and cardiac procedures. We analyzed CHD with an expanded definition to increase statistical power; however, sensitivity analyses also tested more homogeneous outcomes: hard CHD (defined as MI and fatal CHD) and MI. The dataset used for this study includes 3,053 participants with events ascertained up to 2016 (Table 1).

JHS was approved by the IRBs of Jackson State University, Tougaloo College, and the University of Mississippi Medical Center. Informed consent was obtained from all participants.

#### Multi-Ethnic Study of Atherosclerosis (MESA)

The Multi-Ethnic Study of Atherosclerosis (MESA) is a study of the characteristics of subclinical cardiovascular disease and the risk factors that predict progression to clinically overt cardiovascular disease or progression of the subclinical disease^38–40^. MESA consists of a diverse, community-based sample of an initial 6,814 men and women aged 45-84 years without known cardiovascular disease at baseline. Thirty-eight percent of the recruited participants were White, 28 percent African American, 22 percent Hispanic, and 12 percent of Chinese descent. Participants were recruited from six field centers across the United States: Baltimore City and Baltimore County, Maryland; Chicago, Illinois; Forsyth County, North Carolina; Los Angeles County, California; New York, New York; and St. Paul, Minnesota. The first examination took place over two years, from July 2000 to July 2002, and has been followed by additional examinations. The study was approved by the Institutional review boards at all participating institutions, and all participants gave written informed consent. In addition, informed consent was obtained for extensive data sharing (dbGaP) and genetic/omic studies, including candidate genes (NHLBI CARe), genome-wide scans (NHLBI SHARe), exome sequencing (NHLBI ESP) and, most recently, the NHLBI TOPMed program. Our analyses include 870 participants with available DNAm data from exam 1 (Table 1).

Physician-adjudicated CHD events were identified over follow-up through 2019 for the utilized dataset. CHD events were defined as MI, resuscitated cardiac arrest, definite angina, probable angina (if followed by revascularization), and CHD death. As with the JHS, a more expanded definition of CHD was utilized to increase statistical power, followed by sensitivity analyses using hard CHD (MI and fatal CHD) and MI only as more homogeneous outcomes.

#### Methylation Risk Score construction and application

To capture the effects stress across the epigenome, we constructed methylation risk scores (MRSs). We first calculated the coefficients capturing the association between stressful life events (SLEs) and DNAm levels in the WHI using a mixed-effects linear regression model. Given the heterogeneity in study design and available covariates across the contributing WHI ancillary studies, EWAS were conducted separately within WHI-EMPC, WHI-BAA23, and WHI-AS311. Furthermore, because WHI-BAA23 and WHI-AS311 included participants from both the WHI Observational Study and Clinical Trial components, these subsets were analyzed separately to allow adjustment for component-specific covariates and study design factors. Technical factors were modeled as random effects, and each analysis was adjusted for the appropriate study- and component-specific covariates. The resulting effect estimates and standard errors from each stratum were subsequently combined using inverse-variance weighted meta- analysis to obtain overall WHI estimates. Statistically significant CpGs were defined using either a False Discovery Rate (FDR) q-value < 0.05 or a more conservative Bonferroni-corrected p-value < 0.05, yielding 841 and 13 genome-wide significant CpGs, respectively. These meta-analyzed effect estimates were then used as weights for construction of the MRSs, following standard procedures^61,62^, to capture the impact of stress exposure across the epigenome.

Specifically, MRSs were derived by summing the methylation levels, weighted by their stress-related coefficients, of significant CpGs across the epigenome. Using this procedure, two scores were constructed: a more comprehensive score incorporating all 841 FDR-significant CpG sites (MRS_841_) and a more parsimonious score using only the 13 Bonferroni-significant sites (MRS_13_).

To evaluate the association between MRSs and incident CHD, we fitted Cox proportional hazards regression models. However, given the limited number of CHD events and the differences in covariate availability and study design across WHI strata, Cox models were performed by first pooling participants from all ancillary studies into a single analytic cohort and residualizing each MRS on the appropriate stratum-specific covariates. The residualized MRSs were subsequently dichotomized using the cohort median as the cutoff, classifying participants into low- and high-MRS groups, and used as the primary exposure in Cox proportional hazards models. The models included analytic stratum (WHI-EMPC, WHI-BAA23-OS, WHI-BAA23-CT, WHI-AS311-OS, and WHI-AS311-CT) as a covariate to account for residual differences across studies and study components, with incident CHD as the outcome.

The stress-DNAm coefficients trained in the WHI were applied to DNAm levels in the JHS and MESA cohorts to compute MRSs in these independent datasets. MRS values were then adjusted for covariates and evaluated for risk prediction. The WHI cohort was analyzed separately as a cohort for discovery and training, while JHS and MESA were meta-analyzed to independently validate and test generalizability. Given the differences in DNAm platforms used across cohorts, 770 of the 841 FDR-significant stress-related CpGs were available after preprocessing in the JHS EPIC version 1 data, and 773 were available in the JHS EPIC version 2 data. 784 of the 841 sites were present in MESA. Of the 13 Bonferroni-significant CpGs, 12 were present in the JHS (EPIC versions 1 and 2) and MESA.

To confirm that observed associations between MRSs and CHD were specifically driven by our stress-related CpG sites, we conducted 10,000 permutation tests to compare the performance of the stress-related DNAm sites to the whole genome as a background. In each permutation, we randomly selected either 841 or 13 CpG sites—matching the number in MRS_841_ and MRS_13_, respectively—and assigned them the originally trained MRS weights. These permuted scores were residualized for testing as previously described.

#### Functional gene enrichment analysis

Functional enrichment analysis was conducted on the 841 FDR-significant sites using the Genomic Regions Enrichment of Annotations Tool (GREAT^41^). Gene regulatory domains were assigned to each gene according to default parameters. These domains consisted of basal domains up to 5.0 kb upstream and 1.0 kb downstream from the gene’s transcription start site (TSS) and 1.0 Mb extensions up to the basal regulatory domains of genes upstream or downstream of the TSS. The fraction of the genome annotated for an ontology term was calculated against the whole genome background. We selected the top 20 terms that were FDR-significant for both binomial and hypergeometric tests, with a binomial fold enrichment (observed regions hit / expected regions hit) greater than 2.

#### Cell-type-specific DNAm deconvolution

Given that whole blood comprises a heterogeneous mixture of cell types that contribute to bulk-level DNAm signals, we used tensor composition analysis (TCA)^42,43^ to estimate cell-type-specific DNAm levels in association with SLEs in the WHI. Estimates of cell-type proportions for six immune cell types—monocytes, natural killer (NK) cells, granulocytes, CD8 T cells, CD4 T cells, and B cells—were produced using the Horvath online calculator^44,45^. Bulk DNAm data and estimated cell-type proportions were then used as input for TCA to estimate cell-type-specific DNAm levels. TCA models cell-type-specific methylation data as a 3-dimensional tensor of samples by methylation by cell type, in contrast to previous methods which modeled this data as a 2-dimensional matrix of cell type proportions for each sample. Bulk-level DNAm is represented as weighted sums of cell-type-specific parameters, generated using a maximum likelihood- based procedure^42^.

#### Statistical Analyses

All statistical analyses were performed using R versions 4.4.1 and 4.5.0^46^. For all analyses and calculations involving site-level DNAm, beta values representing percent methylation on a scale of 0 (no methylation) to 1 (complete methylation) were used. The discovery EWAS was conducted using linear mixed effects models with the “lme4” function, and stress-MRS analyses were subsequently conducted using “lm” in base R. Meta-analyses were conducted using the “rma” function from the “metafor” package^47^. Figures were created using “ggplot2”^48^. To estimate associations with CHD, Cox proportional hazards regression was conducted on residualized variables (MRSs, single-site DNAm level), wherein positive associations correspond to increased CHD risk. Predictor variables were residualized to prevent Cox regression model instability resulting from low event counts relative to the number of covariates. Cox regression was performed using the “Surv” function from the “survival” package^49^, followed by meta-analyses using “meta.summaries” from the “rmeta” package^50^. Kaplan-Meier curves were created using “survminer”^51^ and “ggplot2”^48^. Participants with a prior history of CHD were excluded from survival analysis. For all meta-analyses, fixed- and random-effects meta-analyses were conducted based on p < 0.05 for heterogeneity.

For all analyses, we adjusted for relevant sociodemographic variables including age, ethnicity, race, education, income, marital status, and neighborhood socioeconomic status calculated as composite scores based on census tract characteristics such as income, wealth, education, and occupation^52–54^. Given that immune cell types exhibit distinct epigenetic signatures and their distribution in blood can be influenced by stress^52^, we also estimated blood cell-type proportions (CD8+ T cells, CD4+ T cells, B cells, NK cells, granulocytes, and monocytes)^44^ from the array DNAm data and included these estimates as covariates in all models using bulk DNAm data. To further adjust for population structure and technical variation, we included the top 10 genetic ancestry principal components and batch effects (plate, well, chip). Models estimating CHD risk additionally adjusted for lifetime smoking (WHI) or current smoking status (JHS and MESA), BMI, and alcohol consumption, known cardiovascular risk factors that can be influenced by stress. Specific to the JHS and MESA, which included both males and females, models were also adjusted for sex.

Prior to mediation analyses using WHI data, MRSs were residualized by regressing on the relevant covariates and subsequently dichotomized using a median split to classify participants into lower- and higher-risk groups. These dichotomized MRS residuals were used as mediators in all mediation models. The mediator model estimated the association between SLEs and the MRS using linear regression. The outcome model estimated the association of SLEs and the MRS with incident CHD using a parametric survival regression model implemented with “survreg”^49^. This approach enabled evaluation of the indirect effect of SLEs on CHD through DNAm variation captured by MRS. Mediation effects were estimated within the counterfactual framework using the *mediation* package (version 4.5.0) in R^55^. Because survival regression follows an accelerated failure time (AFT) framework, the estimated effects represent differences in days to CHD event rather than hazard ratios, wherein positive associations correspond to reduced CHD risk. The average causal mediation effect (ACME), average direct effect (ADE), and proportion mediated were estimated using simulation-based inference. Statistical inference was based on nonparametric bootstrap resampling with 10,000 simulations, and robust standard errors were applied. Confidence intervals and p-values were derived from the bootstrap distributions. All analyses were conducted using complete cases for the exposure, mediator, and outcome variables.

## Results

### 1. Stress burden is associated with genome-wide DNAm differences in the WHI

Our EWAS identified 841 CpG sites significantly associated with SLE burden after FDR correction, and 13 sites remained significant after Bonferroni correction (Fig. 1A). The Q-Q plot showed modest inflation (λ = 1.145), with divergence from the null expectation occurring primarily in the tail of the distribution, supporting the presence of true SLE-associated methylation signals (Fig. S1). Among the 841 significant sites, 99 (11.8%) were hypermethylated and 742 (88%) were hypomethylated (Table S1). Among the 13 Bonferroni-significant CpGs, only two were juxtaposed (within 1 Mb of each other), and these sites were not highly correlated (r < 0.8), indicating that the score likely comprised independent DNAm signals. Top signals for Bonferroni-significant sites include *SRPK2*, which encodes a splicing regulator, and *TNF*, a well-known inflammation-related gene. Of note, Bonferroni genes with known relevance to cardiovascular pathology were identified, including the gene encoding *DOCK10*, which regulates *JNK/p38* signaling and cardiac function under neurohormonal stress^56^, and the gene encoding *MPO*, a neutrophil-derived CVD biomarker related to inflammation and oxidative stress^57^. Notable FDR-significant sites include *ALDH2*, a gene encoding a mitochondrial enzyme involved in alcohol metabolism^58^, *HDAC4*, a gene coding for a histone acetylase involved in epigenetic regulation^59^, and *SLPI*, a circulating leukocyte serine protease inhibitor that regulates inflammation and has been implicated in heart failure in MESA^60^.

**Fig. 1.**
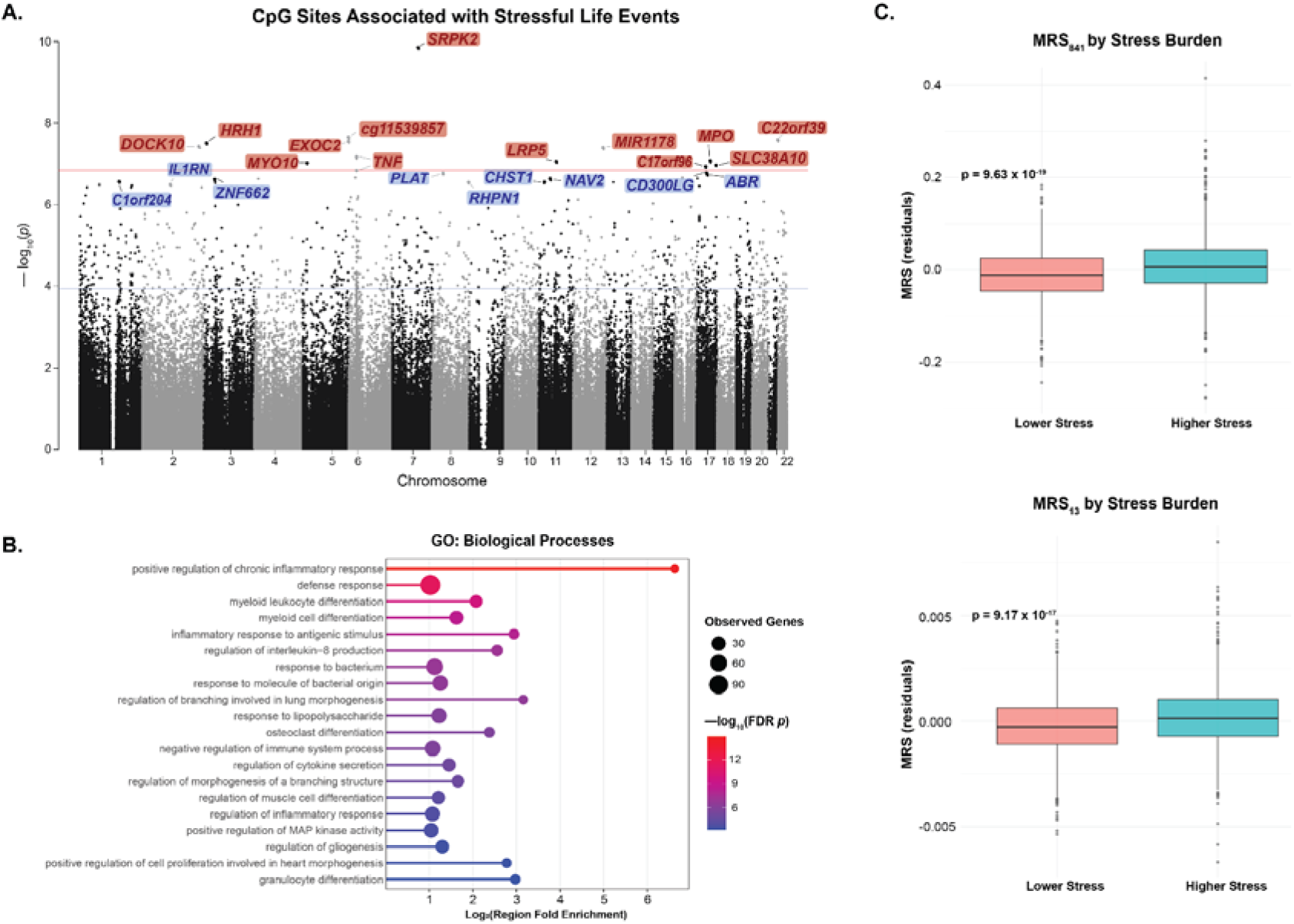
Identification of stress-related CpG sites in the WHI cohort data (n = 3,857). **(A)** Manhattan plot of CpG associations with dichotomized SLE burden from EWAS. SLE burden was significant associated with 841 sites after FDR correction (blue line) and 13 sites after Bonferroni correction (red line). Several of the top genes are labeled blue for FDR-corrected significance and red for Bonferroni-corrected significance. **(B)** Dot plot showing association between SLE-related CpG sites and immune pathways from gene set enrichment analysis. **(C)** Boxplots showing association of stress (stressful life events) burden with MRS₈₄₁ and MRS₁₃ from linear regression analysis.

Gene functional enrichment analysis showed a significant association of stress-related sites with immune-related processes. After FDR correction, differential methylation of the 841 SLE-related sites was associated with regulation of chronic inflammatory response, regulation of cytokine production, and myeloid and granulocyte cell differentiation, among other processes (Fig. 1B).

### 2. Stress-related Methylation Risk Scores predict incident CHD in the WHI

Individuals in WHI with higher SLE burden had significantly higher MRS values compared to those with lower burden, which reflects how the scores were trained in the WHI (MRS_841_: t = 8.9, p = 9.63 x 10^-19^; MRS_13_: t = 8.4, p = 9.17 x 10^-17^; Fig. 1C).

Using Cox proportional hazards regression, we found that each MRS was significantly associated with incident CHD, so that higher MRS (reflecting higher stress burden) is linked to a higher risk of developing CHD over time after adjusting for known risk factors (MRS_841_: Hazard Ratio [HR] = 1.40, 95% Confidence Interval [CI], 1.19 to 1.64, p = 0.0006; MRS_13_: HR = 1.36, 95% CI, 1.15 to 1.60, p = 0.0002) (Fig. 2A & B). Filtering nearby highly correlated CpGs had minimal impact on the MRS_841_, with predictions remaining significant after pruning based on distance cutoffs for 1 kb (819 CpGs; *p* = 0.00038), 10 kb ( 811 CpGs; *p* = 0.00052), and 1 Mb (747 CpGs; *p* = 0.0011). The same relationship was also observed with myocardial infarction (MI) tested as a more homogeneous cardiovascular outcome (MRS_841_: HR = 1.41, 95% CI, 1.17 to 1.71, p = 0.0004; MRS_13_: HR = 1.33, 95% CI, 1.10, 1.61, p = 0.003) (Fig. S2A & B).

**Fig. 2.**
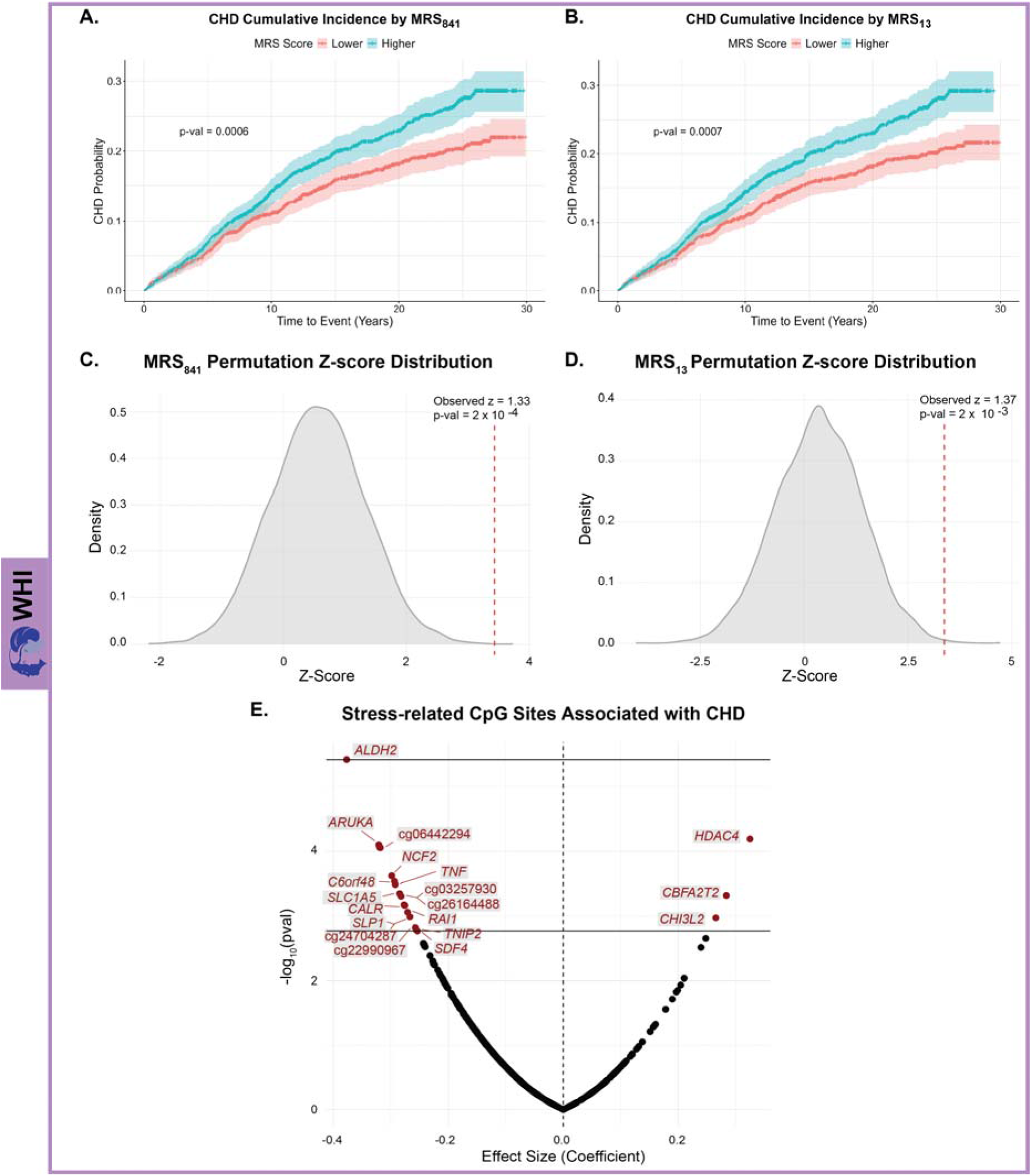
Stress-related MRSs significantly predict CHD risk in the WHI (n = 3,857). (A, **B)** Kaplan-Meier curve showing cumulative CHD incidence by **(A)** MRS₈₄₁ and **(B)** MRS₁₃ level (higher versus lower). **(C, D)** Comparison of CHD prediction by **(C)** MRS₈₄₁ and **(D)** MRS₁₃ (dotted lines) with the distribution of prediction z-scores calculated from array-covered CpG sites selected randomly across 10,000 permutations (continuous lines). **(E)** Volcano plot showing the prediction of CHD by specific stress-related CpG sites at the FDR and Bonferroni levels of statistical significance.

Both MRS_841_ and MRS_13_ significantly outperformed their respective permutation distributions (*P*_perm_ = 0.0002 and 0.002, respectively), indicating that the prediction is unlikely to be due to chance or driven by global DNAm shifts (Fig. 2C & D).

To identify which specific MRS CpGs were most associated with CHD risk, we ran Cox regression models for each site individually. Nineteen CpGs remained significant after FDR correction, and only one site—located on *ALDH2*—remained significant after Bonferroni correction (Fig. 2E, Table S2). *ALDH2* codes for a mitochondrial enzyme that has been previously linked to increased risk for cardiovascular diseases^61,62^. Other notable FDR-significant sites were annotated at *HDAC4* and *TNF,* both of which have been previously associated with cardiovascular function and disease^63,64^.

### 3. MRS and stress-related DNAm sites partially mediate the effect of stress burden on CHD risk

Individuals with higher stress burden had a significantly higher risk of developing CHD over time (HR = 1.23, 95% CI, 1.04 to 1.46, p = 0.016; Fig. 3A). A similar association was found for MI, with increased incidence among those exposed to greater stress burden (HR = 1.23, 95% CI, 1.01 to 1.50, p = 0.038; Fig. S3).

**Fig. 3.**
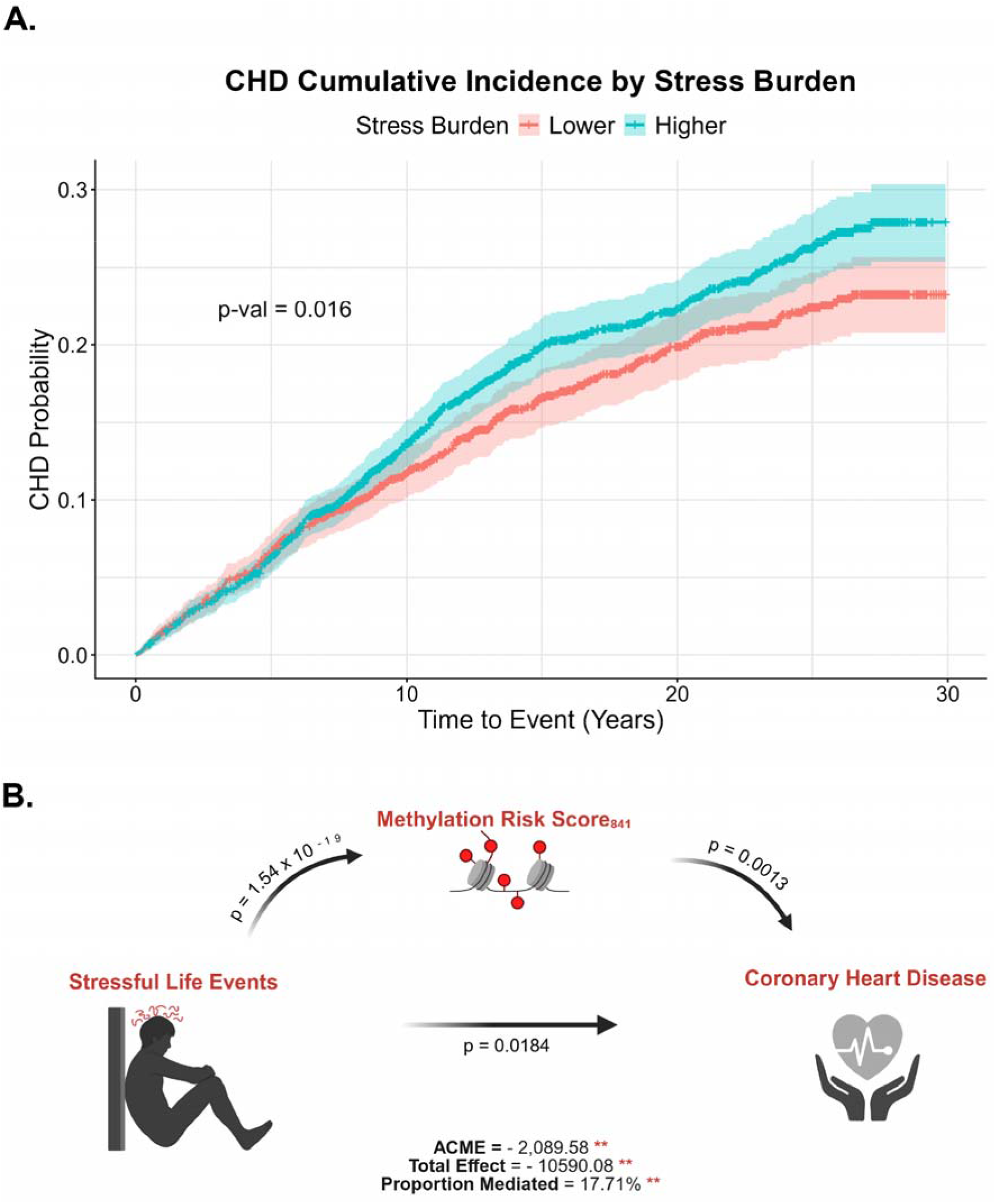
Higher stress (stressful life events) burden is associated with increased CHD risk in the WHI (n = 3,857), mediated by a stress-related methylation risk score (MRS). **(A)** Kaplan-Meier curve showing cumulative CHD incidence by SLE burden level (higher versus lower). **(B)** Schematic showing the relationship between stress burden and CHD with MRS_841_ as a mediator. MRS_841_ statistically significantly mediates the association between SLE and CHD, accounting for 17.71% of the total effect.

In causal mediation analysis, MRS_841_ significantly mediated the association between SLE burden and CHD, with ACME and ADE values reflecting an AFT model in days, wherein negative values correspond to increased risk (ACME = −2,089.58, 95% CI, −3,691.59 to −774.76, p = 0.002). This accounted for approximately 17.71% of the total effect. The direct effect of stress on CHD remained significant (ADE = −8,932.57, 95% CI, −16,600 to –1,394.92, p = 0.018), indicating partial mediation (Fig. 3B). MRS_13_ also significantly mediated the association between SLE burden and CHD (ACME = −1,975.53, 95% CI, −3,496.12 to −699.63, p = 0.001), accounting for approximately 16.49% of the total effect. The direct effect of stress on CHD again remained significant (ADE = −9,094.40, 95% CI, −17,000 to –1,435.50, p = 0.018).

We next determine which of these CpGs were the primary drivers of the mediation effects by testing each of the 841 sites individually. Following FDR correction, 31 sites showed a significant mediation effect, and after Bonferroni correction, only four remained significant (Table S3). Of these four CpG sites, only two could be annotated to a gene: *AURKA* (ACME = -2,541.7, p < 0.001), accounting for approximately 21.8% of the total effect; *and RPL3* (ACME = -1,891.1, p < 0.001), accounting for 16.7% of the total effect. The direct effect of stress on CHD was not statistically significant for either site, suggesting that DNAm at these loci may fully mediate the relationship between stress exposure and CHD risk.

### 4. The prediction of CHD by stress-related MRSs generalizes to the JHS and MESA cohorts

In the generalization cohorts (JHS n = 3,053, MESA n = 870; total n = 3,923), MRS_13_ significantly predicted incident CHD (HR = 1.34, 95% CI, 1.07 to 1.61, p = 0.036), whereas MRS_841_ showed a similar positive association that did not reach statistical significance (95% CI, 1.00 to 1.53, p = 0.087) (Figure 4A & B). These effects did not vary by sex group (MRS_841_ interaction p = 0.66; MRS_13_ interaction p = 0.45). The observed MRS_841_ and MRS_13_ significantly outperformed 10,000 rounds of permutation resampling analysis in CHD risk prediction (P_perm_ = 0.0011 and 0.0087, respectively) (Fig. 4C & D).

**Fig. 4.**
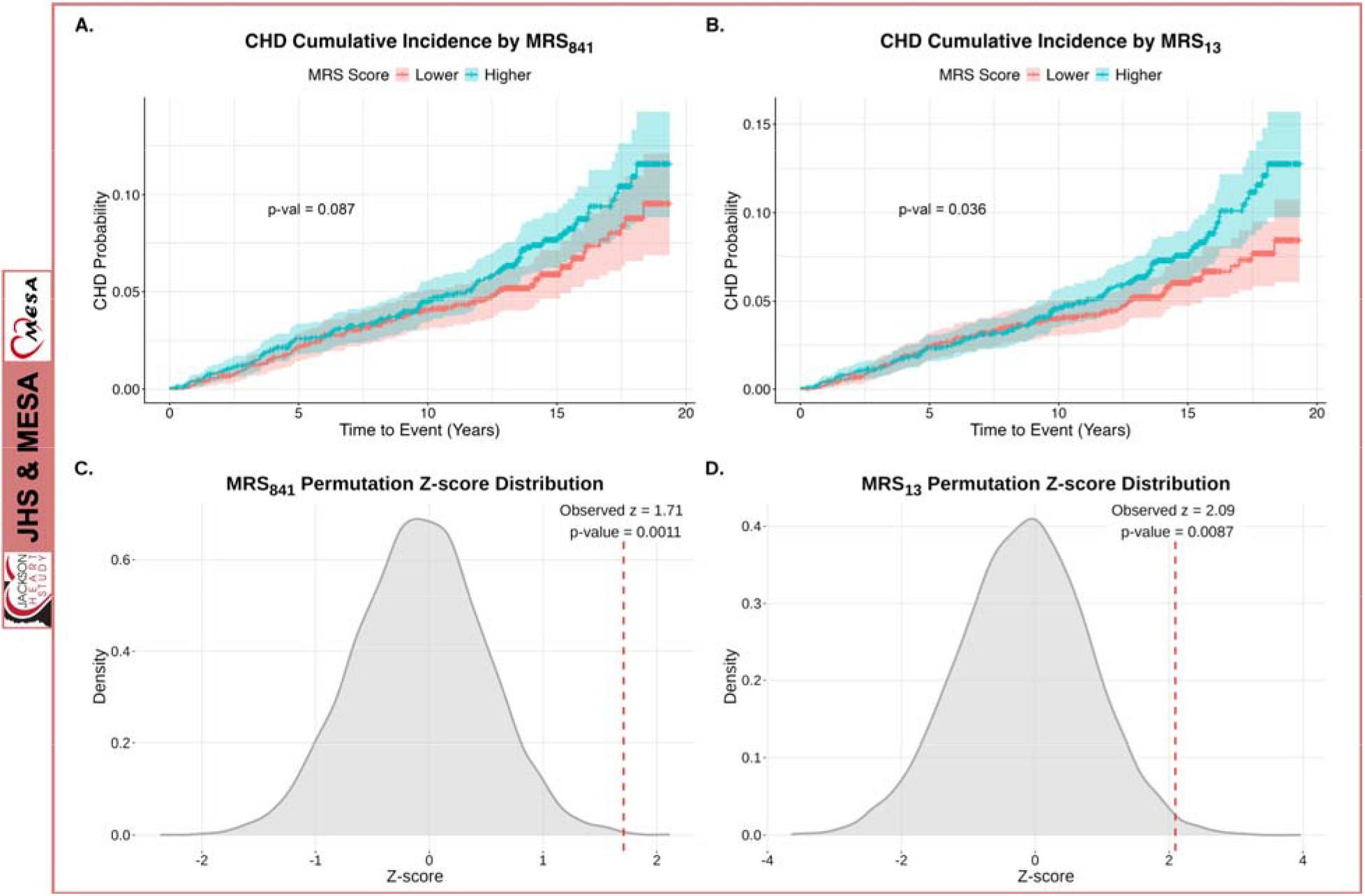
Cox regression and permutation resampling analysis in the validation cohorts (JHS and MESA). (A, **B)** Kaplan-Meier curve showing an association of **(A)** MRS₈₄₁ and **(B)** MRS₁₃ levels with cumulative CHD incidence, meta-analyzed across the JHS and and MESA cohorts (total = 3,923). **(C, D)** Comparison of CHD prediction by **(C)** MRS_841_ and **(D)** MRS_13_ (dotted lines) with the distribution of prediction z-scores calculated from array-covered CpG sites selected randomly across 10,000 permutations (continuous lines).

As an additional sensitivity analysis, we ran Cox regression on the more homogeneous “hard” CHD phenotype (defined as MI and fatal CHD for both the JHS and MESA, with resuscitated cardiac arrest also included for MESA). In line with all CHD event statistics, MRS_13_ significantly predicted hard incident CHD (HR = 1.37, 95% CI, 1.06 to 1.68, p = 0.047), whereas MRS_841_ again showed a similar positive association that did not reach statistical significance (HR = 1.31, 95% CI, 1.00 to 1.61, p = 0.092) (Fig. S4A & B). We also found a positive association between each MRS and incident MI, but this effect did not reach statistical significance (MRS_841_: HR = 1.23, 95% CI, 0.87 to 1.59, p = 0.26; MRS_13_: HR = 1.23, 95% CI, 0.86 to 1.59, p = 0.27) (Fig. S4C & D).

### 5. Cell-type-specific deconvolution of DNAm signals associated with stress burden

While our primary analysis (aiming to identify disease biomarkers) was conducted at the bulk level, we also assessed if specific cell types contributed to the epigenetic signatures of stress. To address this, we applied TCA to deconvolve the bulk DNAm data into cell-type-specific signals in the WHI cohort. Deconvolution was based on DNAm-estimated cell types (CD4+ T cells, CD8+ T cells, monocytes, granulocytes, NK cells, and B cells). Using the 841 CpG sites identified as FDR-significant for their association with stress in the bulk analysis, we subsequently tested whether these associations persisted within each cell type. Among all cell types examined, monocytes exhibited the greatest number of nominally significant stress-related CpG sites that were also predictive of CHD (n = 133), suggesting a prominent role for this cell type in driving the observed associations. Notably, most of the bulk-level CpG sites were hypomethylated in association with stress (88%, 742 out of 841 sites; Fig. 5A). We therefore examined whether this directional pattern was recapitulated at the cell-type level. Monocytes again demonstrated the most pronounced hypomethylation signature among the 841 CpG sites, with 124 sites (93%) showing hypomethylation and only 9 showing hypermethylation (Fig. 5B-C), suggesting that the bulk hypomethylation signal may be driven by monocyte-specific DNAm differences. Conversely, hypermethylated cells, particularly CD8+ T cells (Fig 5C), may counter this bulk hypomethylated signal.

**Fig. 5.**
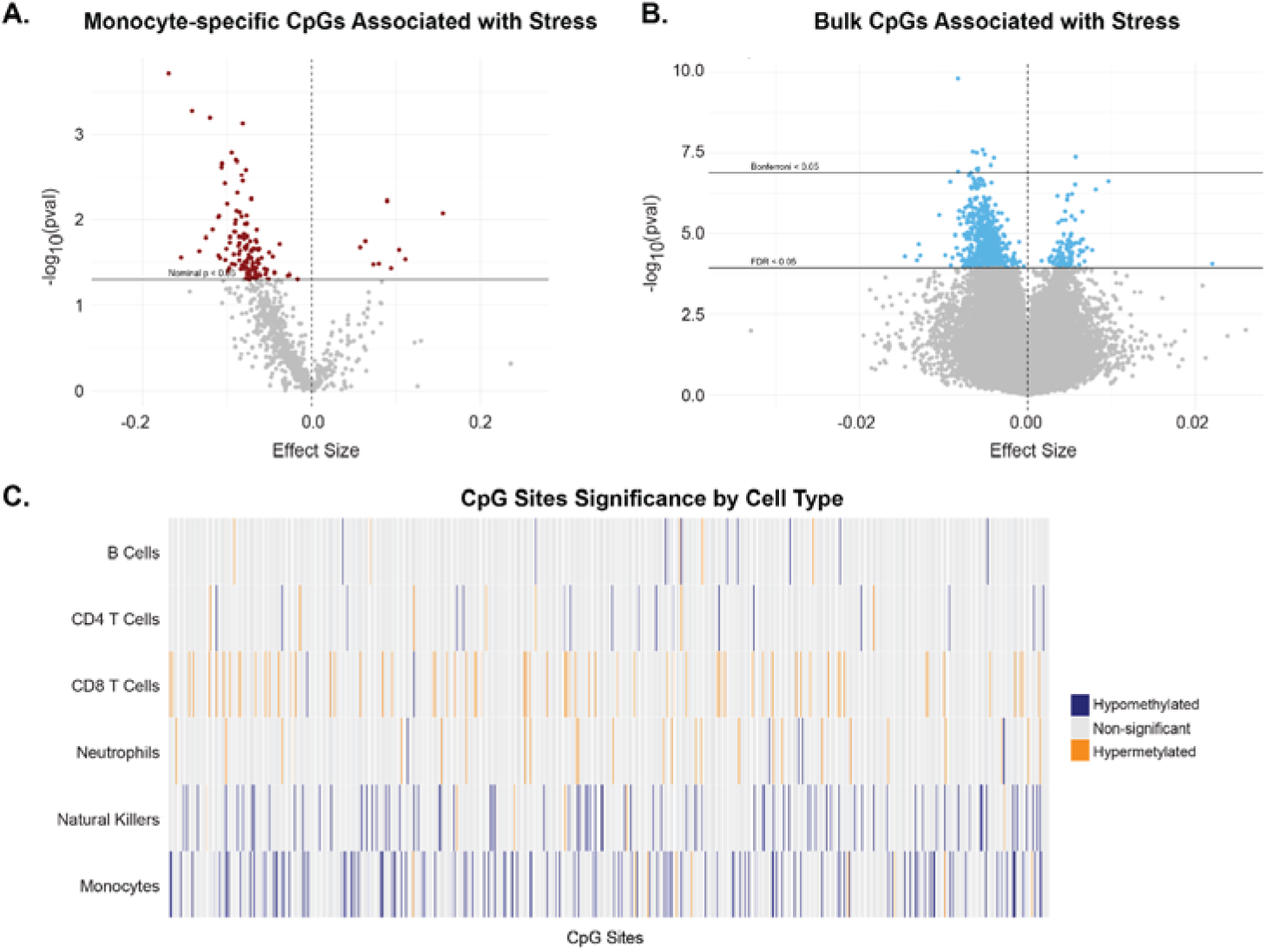
Cell-type DNAm deconvolution analysis reveals cell-type-specific DNAm levels associated with stress burden in the WHI (n = 3,857). (A, **B)** Volcano plots of CpGs associated with SLE burden at the **(A)** monocyte-specific level and **(B)** bulk level for comparison. **(C)** Heatmap of the 841 stress-related CpG sites (ordered by bulk-level significance with lower p-values on the left) by cell type. Monocytes show the most nominally significant differentially methylated sites.

## Discussion

While prior work has associated stress burden with CHD risk^11^, the mechanisms underlying this association are poorly understood. Leveraging three diverse discovery and validation cohorts participating in the TOPMed program, the present study supports methylation risk scores as novel biomarkers of stress-related CHD and further suggests DNAm differences specifically in monocytes as a key link between stress and CHD.

Our discovery analyses focused on the WHI, a cohort of postmenopausal women, since this demographic group experiences elevated stress burden^15–17^ and cardiovascular disease risk^18–20^. Our EWAS in the WHI identified 841 FDR-significant CpG sites associated with SLE burden. Most of these sites (742 or 88%) were found to be hypomethylated in association with stress, in line with previous observations that exposure to stressors and glucocorticoid stress hormones decreases DNAm levels in susceptible genomic regions^25,65,66^. Intriguingly, nine of these sites are located across the gene body, promoter, and 3’-UTR regions of the *TNF* (tumor necrosis factor) gene, which encodes a major proinflammatory cytokine implicated in CHD^64,67^. Circulating *TNF* has been shown to increase in response to acute psychological stress^68^. Moreover, we previously found that decreased *TNF* methylation is associated with blunted negative feedback of the hypothalamic-pituitary-adrenal (HPA) axis and higher ex vivo *TNF* expression following immune stimulation^69^, suggesting *TNF* methylation as a functional signature of stress exposure. Other notable genes identified by our EWAS include *SRPK2* (serine/arginine-rich protein-specific kinase 2), which is involved in splicing and cell cycle regulation^70^, *HDAC4* (histone deacetylase 4), which plays a major role in epigenetic regulation of gene transcription^71^, *DOCK10* (dedicator of cytokinesis 10), a regulator of stress-activated protein kinase pathways^56,72^, *MPO* (myeloperoxidase), a protein released by activated neutrophils during inflammatory processes and marker of atherosclerotic disease^57,73^, and the serine protease inhibitor *SLPI* (secretory leukocyte protease inhibitor), an inflammation regulator that has been associated with heart failure^60,74^. Among the stress-related CpG sites, the top site predicting CHD is located at the gene body of *ALDH2* (Aldehyde Dehydrogenase 2), encoding a mitochondrial enzyme that plays a crucial role in alcohol metabolism and has been previously linked to CVD risk^58^. Beyond identifying single CpG sites and genes, our gene set enrichment analysis shows that the 841 sites are significantly associated with immune processes, most notably with positive regulation of the chronic inflammatory response, which can drive CHD by promoting the development, instability, and rupture of atherosclerotic plaques^75^. Taken together, these data suggest that stress can contribute to differential methylation of genes involved in inflammatory, metabolic, and transcriptional regulatory pathways. Of note, many of these differentially methylated genes, such as *SRPK2* and *ALDH2*, are associated with heterogeneous disease outcomes (such as Alzheimer’s Disease and cancer^58,76,77^), suggesting that stress-related DNAm patterns may potentially contribute to the development of additional non-cardiovascular pathologies.

MRSs have been proposed as a powerful marker of disease risk^78–84^. Analogous to polygenic risk scores, MRSs aggregate the effects of differential DNAm across multiple sites in the epigenome, based on the premise that disease arises from coordinated modest changes at numerous loci. Distinct MRSs have been developed to predict disease outcomes including all-cause mortality^79^, post-traumatic stress disorder^80^, and chronic kidney disease^81^. Furthermore, composite epigenomic markers such as epigenetic clocks have been associated with a range of CVD outcomes including CHD^85,86^ as well as with stress^87^. However, a MRS specifically designed to predict stress-related CVD outcomes had not been developed or validated. Our bulk-level EWAS in the WHI cohort prompted us to construct two stress-related composite DNAm markers: MRS_841_, a score integrating all 841 DNAm sites significantly associated with stress after FDR correction; and MRS_13_, a more parsimonious score integrating only the 13 significant sites after Bonferroni correction. Intriguingly, these markers predicted future incident CHD independent of known risk factors across WHI, JHS, and MESA. In generalization cohorts, MRS_13_ significantly predicted hard and all CHD risk, whereas MRS_841_ showed a positive, albeit statistically nonsignificant, association with hard and all CHD risk. Permutation resampling analyses further supported both MRS_13_ and MRS_841_ as significant predictors of disease relative to randomly permuted MRSs. These findings suggest that stress-related DNAm signatures developed in certain populations (postmenopausal women in the WHI) are generalizable predictors of disease risk in more demographically inclusive populations (men and women in JHS and MESA). In addition to predicting CHD outcomes, both MRSs partially mediated the association between stress and CHD in the WHI cohort, suggesting composite DNAm differences as a potential mechanism underlying this association.

We further implemented TCA to determine which cell types may be contributing to the stress-related DNAm signals. This analysis revealed monocytes as the cell type with the highest number of differentially methylated sites in association with stress (133 out of the 841 sites). Most of these sites (124 or 93%) were hypomethylated, corroborating the bulk-level analyses and suggesting decreased DNAm as a signature of stress exposure in monocytes. These findings extend prior research highlighting the epigenetic impact of stress on monocyte reprogramming^88,89^ and the prominent role of aberrant monocyte function in CVD pathogenesis^90^.

The findings of this study should be interpreted in the context of certain limitations. While many notable SLE-related CpGs were identified and shown to predict CHD, further research is needed to replicate these sites’ association with SLEs in independent cohorts. This replication could not be performed in the JHS and MESA given that these cohorts did not have available SLE burden data at the time of DNAm data collection. For this reason, mediation analysis could not be replicated in these cohorts and should be interpreted with caution. Moreover, experimental approaches are essential to directly establish the downstream functional consequences of SLE-related differential methylation. DNAm of promoter regions is generally associated with reduced gene expression^67^, whereas methylation in other genomic contexts is associated with increased gene expression^91^; however, these trends may not recapitulate across all gene, cell, and tissue types. Dissecting these effects mechanistically can improve understanding of the functional sequelae of differential transcription caused by SLE-related DNAm, ultimately informing biological interpretation and guiding translational applications.

## Supporting information

Data Preprocessing Supplement

## Funding and Acknowledgements

HM was supported in part by a grant from the National Institute of General Medical Sciences under award 5T32 GM135123.

SB was supported in part by a grant from the National Institute of Neurological Disorders and Stroke under award T32NS007431.

Research reported here was supported by the National Heart, Lung, and Blood Institute (NHLBI) of the National Institutes of Health (NIH) under Award Number R01HL163031.

The WHI program is funded by the NHLBI, NIH, and the U.S. Department of Health and Human Services (DHHS) through contracts 75N92021D00001, 75N92021D00002, 75N92021D00003, 75N92021D00004, 75N92021D00005. AS315 was supported by National Institute of Environmental Health Sciences grant R01-ES020836. AS311 was supported by American Cancer Society award 125299-RSG-13-100-01-CCE. BAA23 was supported by NHLBI Broad Agency Announcement contract HHSN268201300006C.

The JHS is supported and conducted in collaboration with Tougaloo College (75N92025D00038), Jackson State University (75N92025D00039), University of Southern Mississippi (75N92025D00040), G.A. Carmichael Family Health Center (75N92025D00041), Wake Forest University Health Sciences (75N92025D00036), and the University of Mississippi Medical Center (75N92025D00037) contracts from the NHLBI with additional support from the National Institute of Minority Health and Health Disparities (NIMHD).

MESA (phs001416.v1.p1) was performed at the Broad Institute of MIT and Harvard (3U54HG003067-13S1). Centralized read mapping and genotype calling, along with variant quality metrics and filtering were provided by the TOPMed Informatics Research Center (3R01HL-117626-02S1). Phenotype harmonization, data management, sample-identity QC, and general study coordination, were provided by the TOPMed Data Coordinating Center (3R01HL-120393-02S1). The MESA projects are conducted and supported by the NHLBI in collaboration with MESA investigators. Support for MESA is provided by contracts 75N92025D00022, 75N92020D00001, HHSN268201500003I, N01-HC-95159, 75N92025D00026, 75N92020D00005, N01-HC-95160, 75N92020D00002, N01-HC-95161, 75N92025D00024, 75N92020D00003, N01-HC- 95162, 75N92025D00027, 75N92020D00006, N01-HC-95163, 75N92025D00025, 75N92020D00004, N01-HC-95164, 75N92025D00028, 75N92020D00007, N01-HC- 95165, N01-HC-95166, N01-HC-95167, N01-HC-95168, N01-HC-95169, UL1-TR- 000040, UL1-TR-001079, UL1-TR-001420, UL1TR001881, DK063491, and R01HL105756. The authors thank the MESA participants and the MESA investigators and staff for their valuable contributions. A full list of participating MESA investigators and institutions can be found at http://www.mesa-nhlbi.org

Molecular data for the Trans-Omics in Precision Medicine (TOPMed) program was supported by the NHLBI. Genome sequencing for “NHLBI TOPMed: The Jackson Heart Study” (phs000964.v1.p1) was performed at the Northwest Genomics Center (HHSN268201100037C, HHSN268201600032I). Genome sequencing for “NHLBI TOPMed: MESA and MESA Family AA-CAC” (phs001416) was performed at the Broad Institute (HHSN268201600034I, 3U54HG003067-13S1). Methylation for “NHLBI TOPMed: MESA and MESA Family AA-CAC” (phs001416) was performed at the <u>KeckMolecular Genomics Core Facility</u> (HHSN268201600034I). Core support including centralized genomic read mapping and genotype calling, along with variant quality metrics and filtering were provided by the TOPMed Informatics Research Center (3R01HL-117626-02S1; contract HHSN268201800002I). Core support including phenotype harmonization, data management, sample-identity QC, and general program coordination were provided by the TOPMed Data Coordinating Center (R01HL-120393; U01HL-120393; contract HHSN268201800001I). We gratefully acknowledge the studies and participants who provided biological samples and data for TOPMed.

## Data Availability

The code used for all analyses are available on Github. All data produced are contained in the manuscript or available from the authors upon reasonable request.

https://github.com/hazelmilla/MRS_TOPMed.git

