## Supplemental Figures for "Stress-Related Methylation Risk Scores Predict Coronary Heart Disease"

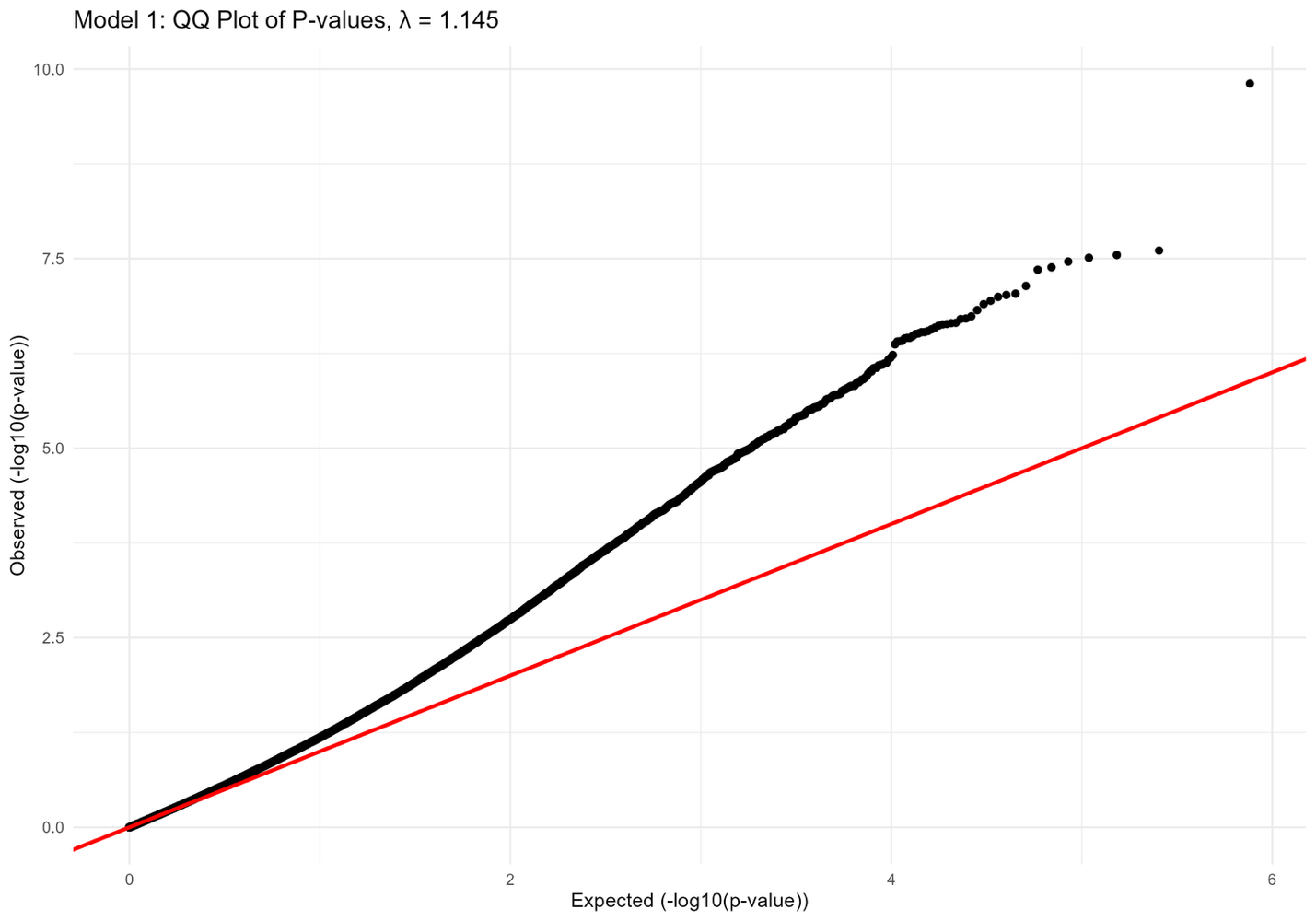

**Fig. S1.** Q-Q plot of observed p-values.

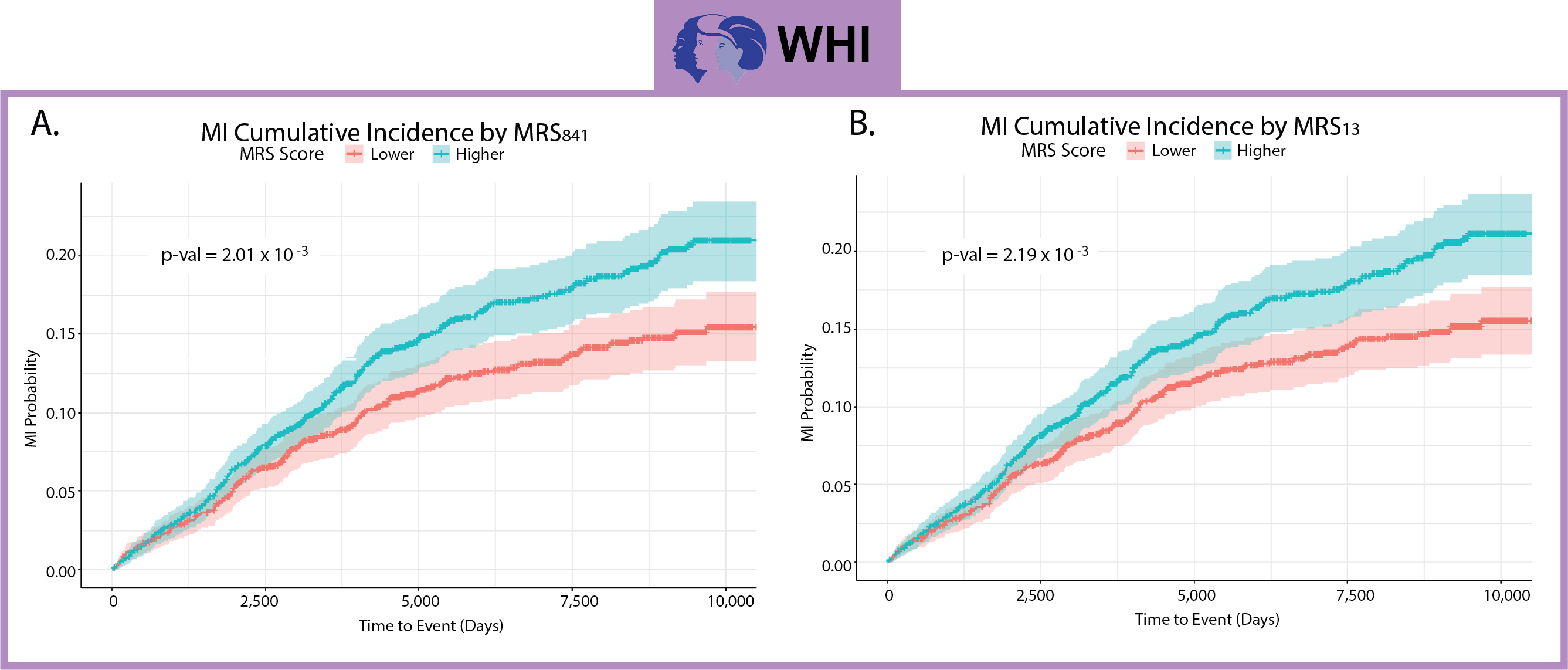

**Fig. S2.** Stress-related MRSs significantly predict the risk of myocardial infarction (MI) in the WHI (n = X). (A, B) Kaplan-Meier curve showing cumulative MI incidence by (A) MRS₈₄₁ and (B) MRS₁₃ level (higher versus lower).

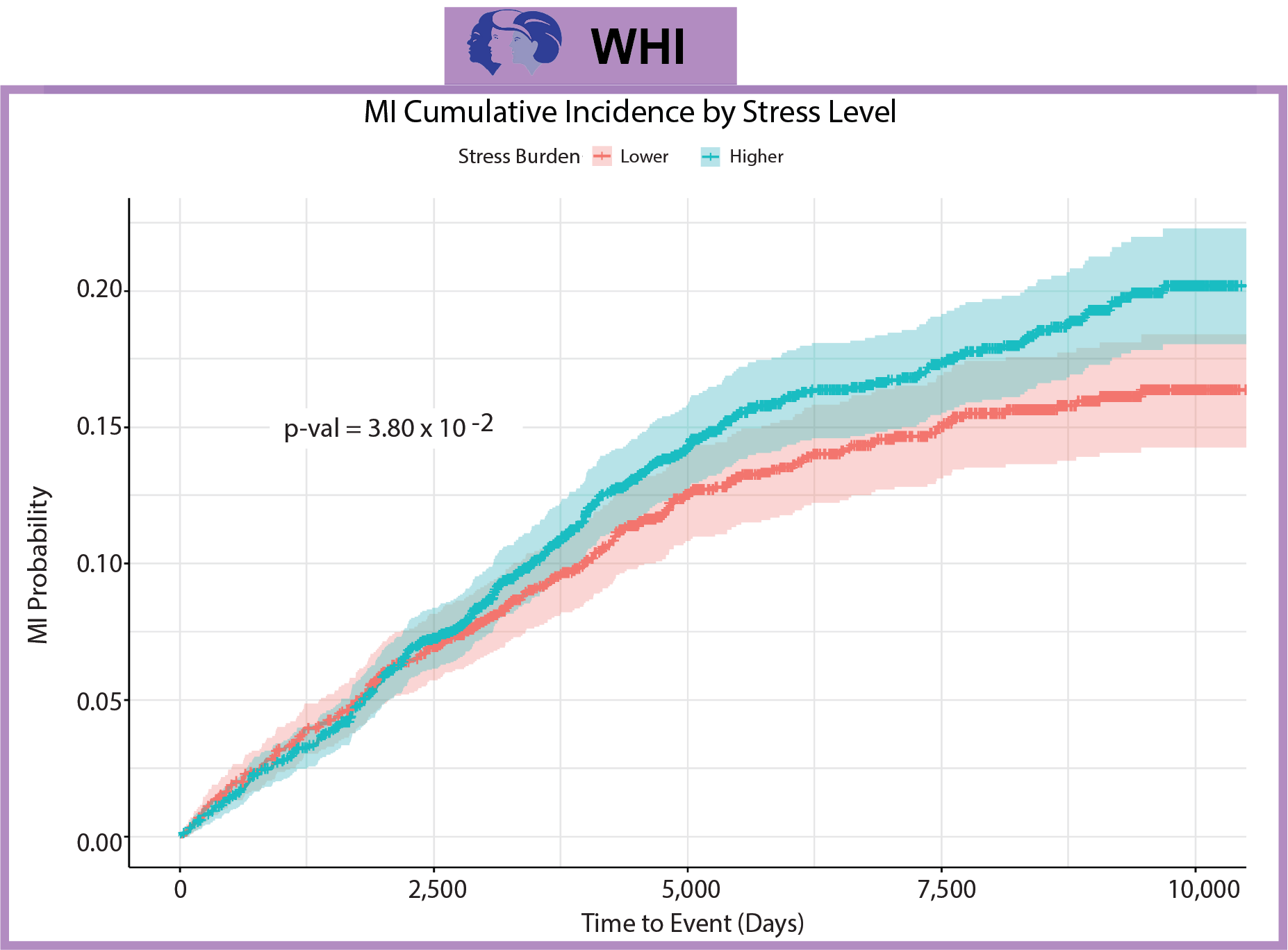

**Fig. S3.** Higher stress (stressful life events) burden is associated with increased MI risk in the WHI (n = 3,857). Kaplan-Meier curve showing cumulative incidence of MI by SLE burden level (higher vs. lower).

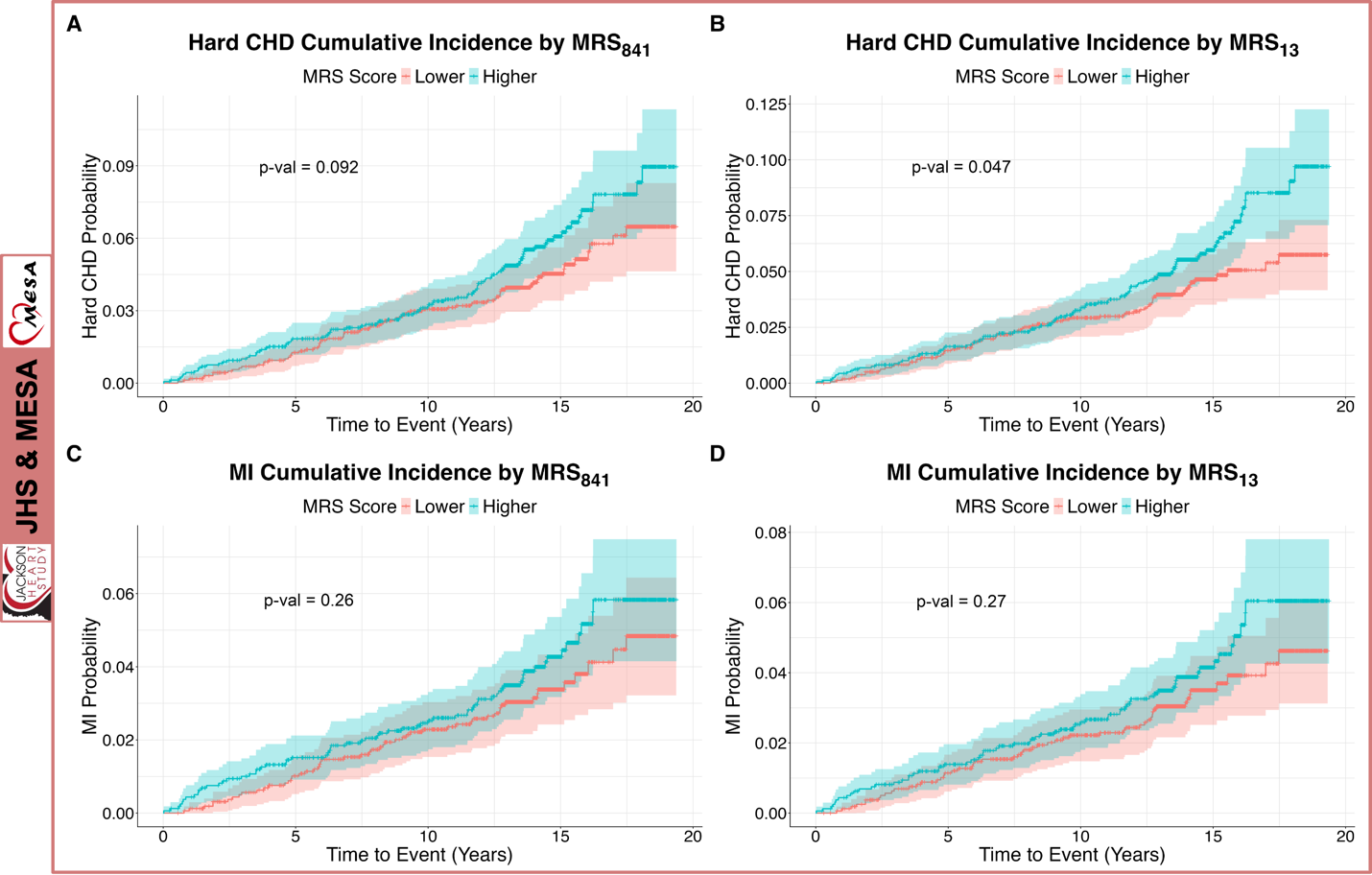

**Fig. S4.** Higher MRS levels are associated with increased hard CHD (A, B) and MI (C, D) risk in the JHS and MESA (n = 3,923).
