## Supplementary material for "Stress-Related Methylation Risk Scores Predict Coronary Heart Disease": Data Preprocessing Supplement

**DNAm processing**

*WHI*

DNA was extracted from whole-blood samples collected at the baseline screening visit and was bisulfite converted using the Zymo EZ DNA Methylation Kit. Genome-wide DNA methylation was assessed using the Illumina Infinium HumanMethylation450 BeadChip (Illumina Inc., San Diego, CA, USA). Quality control, probe filtering, and normalization were performed independently within each study following established protocols^1^. Probe-type bias arising from differences between Infinium I and II chemistries was corrected using Beta Mixture Quantile (BMIQ) normalization^2^. Technical covariates, including plate, chip, row, and column, were available across studies and accounted for in regression models. In the WHI-EMPC, batch effects were additionally corrected using the nonparametric empirical Bayes ComBat method^3^.  DNA methylation levels were quantified as beta values, representing the proportion of methylated cytosines relative to the total methylated and unmethylated signal. Array DNAm data were used to estimate blood cell proportions (CD8+ T cells, CD4+ T cells, B cells, natural killer cells, granulocytes, monocytes^4^) that were included as covariates to adjust for potential confounding in regression models.

*JHS*

DNAm data were collected from whole-blood samples taken at baseline examination and assessed using the Illumina Infinium Methylation EPIC BeadChip Array (Illumina Inc., San Diego, CA). Two versions of the EPIC BeadChip Array were used: version 1 (EPICv1), covering ~850K sites, and version 2 (EPICv2), covering ~930K sites. Datasets from different versions of the EPIC BeadChip array were processed and analyzed separately to account for assay-specific differences. Only probes present in both datasets after QC and analysis were retained for meta-analysis.

We utilized the Chip Analysis Methylation Pipeline (*ChAMP*) to import and filter probes based on signal quality according to default parameters (detection p-value < 0.01)^5,6^. Samples were filtered out if the proportion of low-quality probes exceeded the default threshold value (0.1). Probes with a low bead count (<3 beads in at least 5% of samples per probe) were additionally filtered out. For EPICv1 data, data from one ancillary study (ASN0104) were retained while data from an ancillary study with fewer participants with DNAm data (ASN0148) were removed to avoid study-specific confounders in this dataset.

Beta values were input into the Horvath calculator^7^ for imputation of six blood cell-type proportions (granulocytes, monocytes, natural killer, CD4+ T cells, CD8+ T cells, B cells^4^). Type II probe data were normalized to fit the statistical distribution of type I probes using beta-mixture quantile (BMIQ) dilation^2^; then cross-hybridizing^8^, SNP-associated^9^, and non-CpG probes, as well as the 1000 least variable probes, were removed. ComBat correction^3^ was implemented using the *sva*^10^ and *ChAMP*^5,6^ packages to adjust for technical covariates (plate, well, array). Probes located on the X and Y chromosomes were then removed. For the EPICv2 dataset, duplicate probes were removed at random, as replicate probe signals are highly correlated^11^.

*MESA*

DNAm data were generated from whole blood using the Infinium Methylation EPIC BeadChip version 1 (~850 K CpG sites). Probes were filtered at detection p-value > 0.05 to filter out background signal using the *minfi* package^12,13^. Single sample Noob (ssNoob) was used for normalization^13^ to derive corrected methylated and unmethylated signals for conversion to beta values^14^. Samples were dropped based on SNP non-concordance (n=1) and related pairs (n=5). Six samples were dropped as visual outliers based on principal component data. Probes with poor genomic mapping quality, non-unique 30 bp flanking sequence, and extension base inconsistent with color channel were removed^9^. We additionally filtered out SNP-associated, duplicate, least variable, non-CpG, and XY chromosome probes.
